# Transmission of SARS-CoV-2 in Norwegian schools: A population-wide register-based cohort study on characteristics of the index case and secondary attack rates

**DOI:** 10.1101/2021.10.04.21264496

**Authors:** Torill Alise Rotevatn, Vilde Bergstad Larsen, Tone Kristin Bjordal Johansen, Elisabeth Astrup, Pål Surén, Margrethe Greve-Isdahl, Kjetil Telle

## Abstract

**Objectives:** To assess transmission of SARS-CoV-2 in schools mainly kept open during the COVID-19 pandemic.

**Design:** Population-wide register-based cohort study.

**Setting:** Primary and lower secondary schools in Norway have been open during the academic year 2020/2021 with strict infection prevention and control (IPC) measures in place. All identified contacts including student and staff members were urged to get tested following a positive SARS-CoV-2 case in a school.

**Participants:** All students and educational staff in Norwegian primary and lower secondary schools from August 2020 to June 2021.

**Main outcome measures:** Overall secondary attack rate (SAR14) was operationalized as the number of secondary cases (among students and/or staff) in the school by 14 days after the index case, divided by the number of students and staff members in the school. Moreover, we calculated *SAR14-to-students*, denoting transmission from all index cases to students only, *SAR14-to-school staff*, denoting transmission from all index cases to staff members only. We also calculated these measures in stratified samples consisting of student index cases or school staff index cases.

**Results:** From August 2020 to June 2021 there were 4,078 index cases, 79% were students and 21% were school staff. In the majority (55%) of schools with an index case, no secondary cases were observed by 14 days, and in 16% of the schools there were only one secondary case within 14 days. Overall SAR14 was found to be 0.33% (95%CI 0.32-0.33). Staff-to-staff transmission (SAR14 0.45%, 95%CI 0.40-0.52) was found to be slightly more common than student-to-student (SAR14 0.33%, 95%CI 0.32-0.34) and student-to-staff (SAR14 0.28%, 95%CI 0.25-0.30) transmission.

**Conclusions:** Our results confirm that schools have not been an important arena of transmission of SARS-CoV-2 in Norway and therefore support that schools can be kept open with IPC measures in place.

## Background

The role of schools as arena for transmission of SARS-CoV-2, the virus causing COVID-19, has gained much attention during the pandemic. Although children can contract and transmit the virus, this seems to happen less frequently than in adults (1, 2). Growing evidence shows that risk of transmission between students and staff in schools is low when infection prevention and control (IPC) measures are implemented and adhered to (3). However, most studies on transmission in schools have been conducted when re-opening after a period of school closure, when community transmission is decreasing. The risk of introducing the virus in schools by students and staff increases with higher community incidence (3). Thus, many countries closed schools in periods with rising community infection rates (4, 5). Information on transmission in schools during periods with high community infection rates is therefore sparse, especially in the context of more transmissible virus variants (3).

In Norway, schools were closed for six weeks when the first infection wave hit in the spring of 2020. However, to diminish the negative psychosocial and educational consequences of school closure (2, 6-9), schools were overall kept open during the academic year 2020/2021 (i.e. from August 2020 to June 2021). IPC measures were carefully implemented and adapted to the local epidemiological situation. With increasing incidence, strict measures to enable social distancing and organizing students in small cohorts were implemented. The strictest measures sometimes necessitated part time digital teaching for older students. Face masks were not part of measures in primary schools, and played only a minor role in secondary schools, implemented in late spring 2021 when the alpha wave was declining. Widespread use of testing, isolation, contact tracing and quarantine was additionally used to limit spread in the society (10-12).

Studies of transmission into and within schools are important to improve our understanding of the role of students and school staff for the spread of the virus, and in particular how IPC measures may counteract transmissions between students and school staff. The situation in Norway with very limited use of school closures in primary and secondary schools during the academic year of 2020/2021 establishes an interesting setting for studying transmissions within schools in the presence of presumably well-functioning IPC measures. By using individual level data from administrative registries covering every resident of Norway over time, we analysed index cases and secondary transmissions in all primary and lower secondary schools in Norway during the academic year 2020/2021.

## Methods

### Data sources

In this register-based cohort study, we used data in Beredt C19, an emergency preparedness register developed to provide rapid knowledge on the COVID-19 pandemic in Norway (13). From within Beredt C19, we compiled individual-level information originating from the following administrative registries: demographic information from the National Population Registry; date of positive polymerase chain reaction (PCR) tests for SARS-CoV-2 from the Norwegian Surveillance System for Communicable Diseases (MSIS); date of conducted SARS-CoV-2 PCR test regardless of test result from the MSIS Laboratory Database; employment contracts for school staff members from the Employer- and Employee-register and encrypted school organizational numbers to identify school attendance and employment from the National Education Database from Statistics Norway.

In the 2020/2021 academic year, there were 2,776 registered primary and lower secondary schools in Norway (14). Our data contained information on 2,641 schools and their respective geographical school catchment areas. We linked each student to schools based on their place of residence and birth year to determine student school affiliation (see appendix for details).

### Study population

The study period was set to the academic year 2020/2021, i.e., from August 17^th^, 2020, to June 15^th^, 2021. Our study population consisted of all students in primary and lower secondary school age (primary school students born 2008-2014; lower secondary school students born 2005-2007) and all employees aged 20-70 years in the 2,641 schools, who were registered as Norwegian residents in August 2020. Only employees in occupations that usually imply frequent contact with students (teachers, child-care workers, and teaching assistants, table A-1) were included. The registry data are of high quality and include information of the overall population over time, which enable complete follow-up with no attrition (except emigration and death).

### Index cases, secondary cases, and infection clusters

Within each school, the index case was defined as the first student or staff member with a positive SARS-CoV-2 PCR test. Secondary cases included all non-index students or staff at the same school as the index case with a positive PCR test within 14 days of the index case’s positive test date. Index and secondary cases were clustered together if cases occurred at the same school within a period of 14 days or less into the following categories: *single case clusters, two-case clusters*, and *multiple-case clusters* (>2 cases). All cluster types were considered ended after 14 days without any new cases at the same school, and schools were thereafter re-entered if new cases were detected. No index cases and associated secondary cases were included after May 31^st^, 2021 to ensure complete follow-up.

### Outcome

In accordance with the literature, we used secondary attack rates (SAR) as our measure of onward transmission of SARS-CoV-2 (15, 16). We calculated SAR14 as the number of secondary cases divided by the total number of students and staff members in every school with an index case, multiplied by 100. Index cases were excluded in both the numerator and denominator. Clusters with two or more co-index cases, i.e., index cases with identical test dates, were excluded.

### Statistical analyses

First, to provide an overview of the national infection rates during the study period, we plotted the weekly number of new cases per 100,000 residents between week 34, 2020 and week 23, 2021 in total in Norway. We also calculated the the proportion of detected cases among students and staff in the study population. Second, we stratified index cases by cluster type to study differences in distribution and characteristics. Third, we calculated *overall SAR14*, denoting transmission from all index cases to both students and staff members; *SAR14 to students*, denoting transmission from all index cases to students only; and *SAR14 to school staff*, denoting transmission from all index case to staff members only. In addition, we calculated these measures for student index cases and school staff index cases separately, as well as separately for index cases by occupational group (teachers, teaching assistants, child-care workers). To study changes in trends over time and between periods where different SARS-CoV-2 variants have dominated, we visualized the overall SAR14 by month of positive test date for student and school staff index cases. We also calculated the proportion of tested students and staff at schools within 14 days of the index case to assess the relationship between SAR and testing behavior. As a sensitivity analysis, we calculated SAR14 to students within each student birth cohort, since students were likely to have closer contact with other students at the same age, e.g., as implemented IPC measures involved restriction of close contact between students and staff across grades. In all analyses, 95% confidence intervals were calculated using the Wilson method for binomial proportions. All statistical analyses were performed using R Statistical Software (version 3.6.2).

### Ethical approval

The emergency preparedness register, BEREDT C19, was established according to the Health Preparedness Act §2-4 and the project was approved by the Ethics Committee of South-East Norway (9th March 2021, #198964).

## Results

A total of 118,629 cases of confirmed COVID-19 were registered in MSIS between 17^th^ August 2020 and 15^th^ June 2021. Incidence peaked in week 46, 2020, and in week 1 and week 11, 2021 (figure 1).

**Figure 1.**
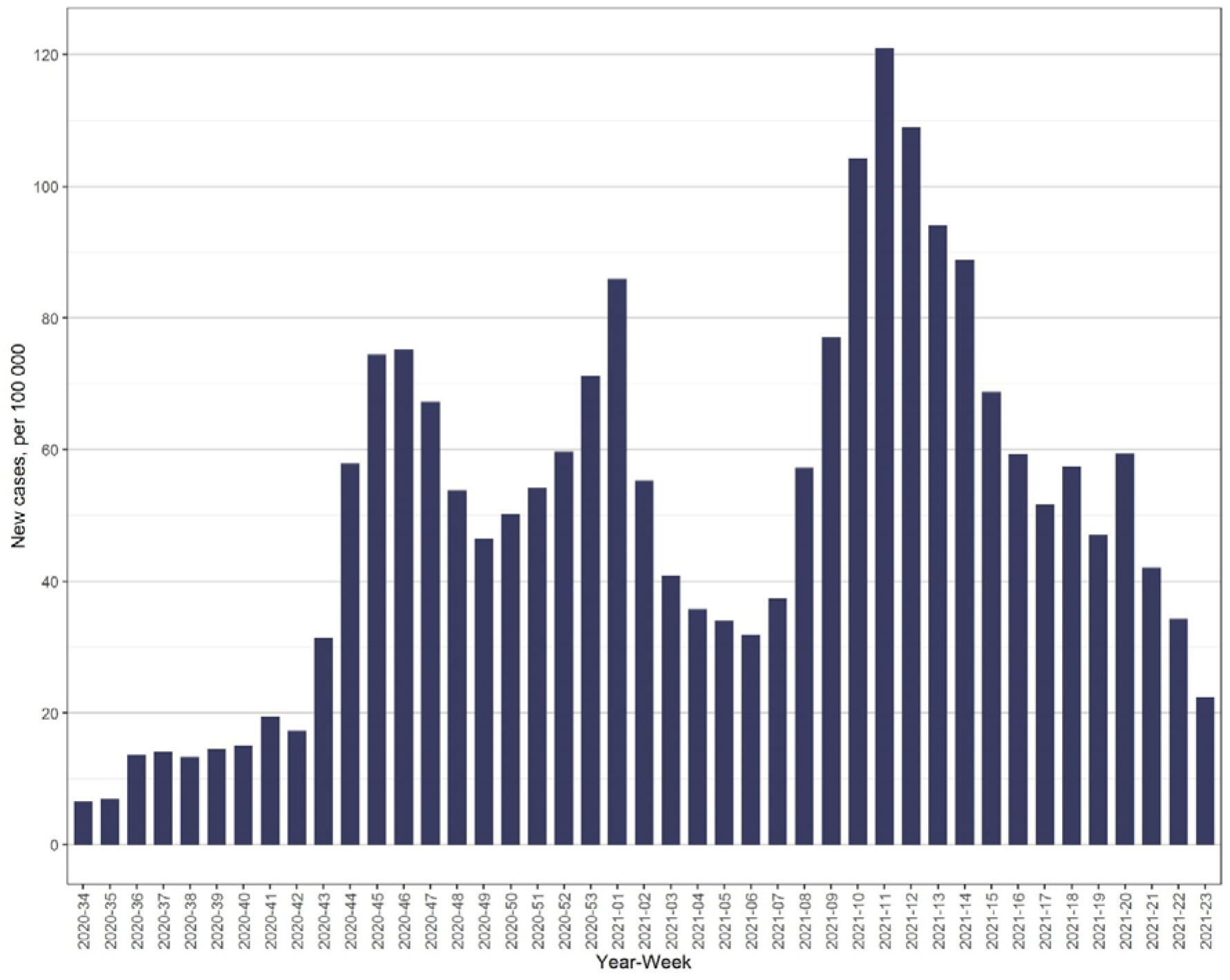
Number of COVID-19 cases per 100.000, per week in Norway in the period between week 34, 2020 and week 23, 2021.

A total of 640,295 students and 102,574 staff members in 2,641 different schools were included in the study population, of which 15,390 (2.4%) students and 2,419 (2.4%) staff members tested positive for COVID-19. There were large geographical variations, with the region of Oslo having highest infection rates among both students (6.2%) and staff members (7.3%; figure A-1). The corresponding proportion for the full Norwegian population during the study period was 2.2%.

### Characteristics of index cases

A total of 586 infection clusters involving 3,609 cases had two or more co-index cases and were therefore excluded from further analysis. This left 12,217 and 2,073 positive student and school staff cases.

In total, we identified 4,078 index cases in 1,573 different schools, of which 79.0% were students and 21.0% were staff members (table A-2). No secondary cases were observed by 14 days in the majority (54.7%) of clusters, and only one secondary case was detected in 15.5% of clusters. Multiple-case clusters were detected in 29.8% of the clusters. Multiple-case clusters were more common in larger schools (<100 students: 10.0%, 100-299 students: 21.2%, 300+ students: 34.8%), while single case clusters were more common in smaller schools (<100 students: 75.6%, 100-299 students: 64.1%, 300+ students: 49.3%). A higher proportion of single case clusters were observed in teachers compared with students (60.8% vs. 53.0%), and a higher proportion of multiple-case clusters were observed in lower secondary schools than in primary schools (32.1% vs 28.4%).

### SAR14 in Norwegian schools

Among all 4,078 index cases, the overall SAR14 was 0.33% (95% CI 0.32-0.33). This means that an average of 0.33% of students and staff had confirmed COVID-19 within 14 days of the index case (table 1), or, put differently, that there were about 3 secondary cases per 1,000 index cases. SAR14 remained similar when restricting secondary cases to students and staff, respectively. Small schools with fewer than 100 students had higher overall SAR14 (0.60%, 95% CI 0.49-0.74) than both medium-sized (0.36%, 95% CI 0.34-0.39) and large (0.32%, 95% CI 0.31-0.32) schools. The same pattern was also observed for SAR14 to students, while school size did not impact SAR14 to staff.

**Table 1:** Secondary attack rates (%) within 14 days after index test date (SAR14) stratified by demographic characteristics of index case.

| Characteristics of index case | Overall SAR14 | SAR14 to students | SAR14 to staff |
| --- | --- | --- | --- |
| **All index cases, all school levels** | **0.33 (0.32-0.33)** | **0.33 (0.32-0.34)** | **0.31 (0.29-0.34)** |
| **School size (all index cases)** |  |  |  |
| < 100 | 0.60 (0.49-0.74) | 0.69 (0.56-0.87) | 0.35 (0.21-0.58) |
| 100-299 | 0.36 (0.34-0.39) | 0.38 (0.35-0.40) | 0.30 (0.25-0.35) |
| 300+ | 0.32 (0.31-0.32) | 0.32 (0.31-0.33) | 0.32 (0.29-0.35) |
| **Students** | **0.32 (0.31-0.33)** | **0.33 (0.32-0.34)** | **0.28 (0.25-0.30)** |
| Primary school age | 0.31 (0.30-0.32) | 0.31 (0.30-0.32) | 0.30 (0.27-0.34) |
| Lower secondary school age | 0.34 (0.32-0.36) | 0.36 (0.34-0.37) | 0.23 (0.20-0.27) |
| **School staff** | **0.34 (0.32-0.36)** | **0.32 (0.30-0.34)** | **0.45 (0.40-0.52)** |
| Primary school | 0.37 (0.35-0.40) | 0.35 (0.32-0.38) | 0.50 (0.43-0.59) |
| Combined schools<sup>b</sup> | 0.25 (0.22-0.29) | 0.23 (0.20-0.27) | 0.36 (0.27-0.49) |
| Lower secondary school | 0.34 (0.30-0.39) | 0.34 (0.29-0.38) | 0.40 (0.29-0.54) |
| Teacher | 0.37 (0.35-0.39) | 0.35 (0.33-0.37) | 0.49 (0.43-0.57) |
| Teaching assistants | 0.25 (0.21-0.29) | 0.24 (0.19-0.29) | 0.31 (0.21-0.47) |
| Child-care workers | 0.23 (0.18-0.29) | 0.21 (0.16-0.27) | 0.35 (0.21-0.57) |

*Notes:*
95 % confidence intervals in parentheses calculated using the Wilson method for binomial proportions. SAR14 was calculated using the number of infected non-index students and staff members in the numerator and the total number of non-index students and staff members in the denominator. <sup>b</sup>Schools where primary and lower secondary school are combined.

When the index case was a student, 0.33% (95% CI 0.32-0.34) of other students and 0.28% (95% CI 0.25-0.30) of staff members had confirmed COVID-19 within 14 days. For index case students in primary school, there was no difference in SAR14 to students and to staff. However, for index case students in lower secondary school, the SAR14 was higher to fellow students 0.36% (95% CI 0.34-0.37) than to staff members (0.23%, 95% CI 0.20-0.27).

When the index case was a staff member, 0.45% (95% CI 0.40-0.52) of the staff members and 0.32% (95% CI 0.30-0.34) of the students were found to have confirmed COVID-19 within 14 days. This suggest that staff-to-staff transmission was higher than staff-to-student transmission, also by separate occupations (teachers, teaching assistants, and child-care workers). The highest transmission rate was found from staff to other staff in primary schools (0.50%, 95% CI 0.43-0.59).

SAR14 from staff index cases to staff or student secondary cases was slightly higher than from student index cases to staff or student secondary cases during the second wave of COVID-19 in Norway, starting in October 2020 (figure 2). This SAR14 peaked in October for staff index cases and in November for student index cases. From January 2021, around the beginning of the third wave, there was a relatively steep increase in SAR14 for both student and staff index cases, followed by a second peak in both groups in March. From February onwards when the Alpha virus variant became dominant, there was no significant difference in SAR14 between student and staff index cases.

**Figure 2:**
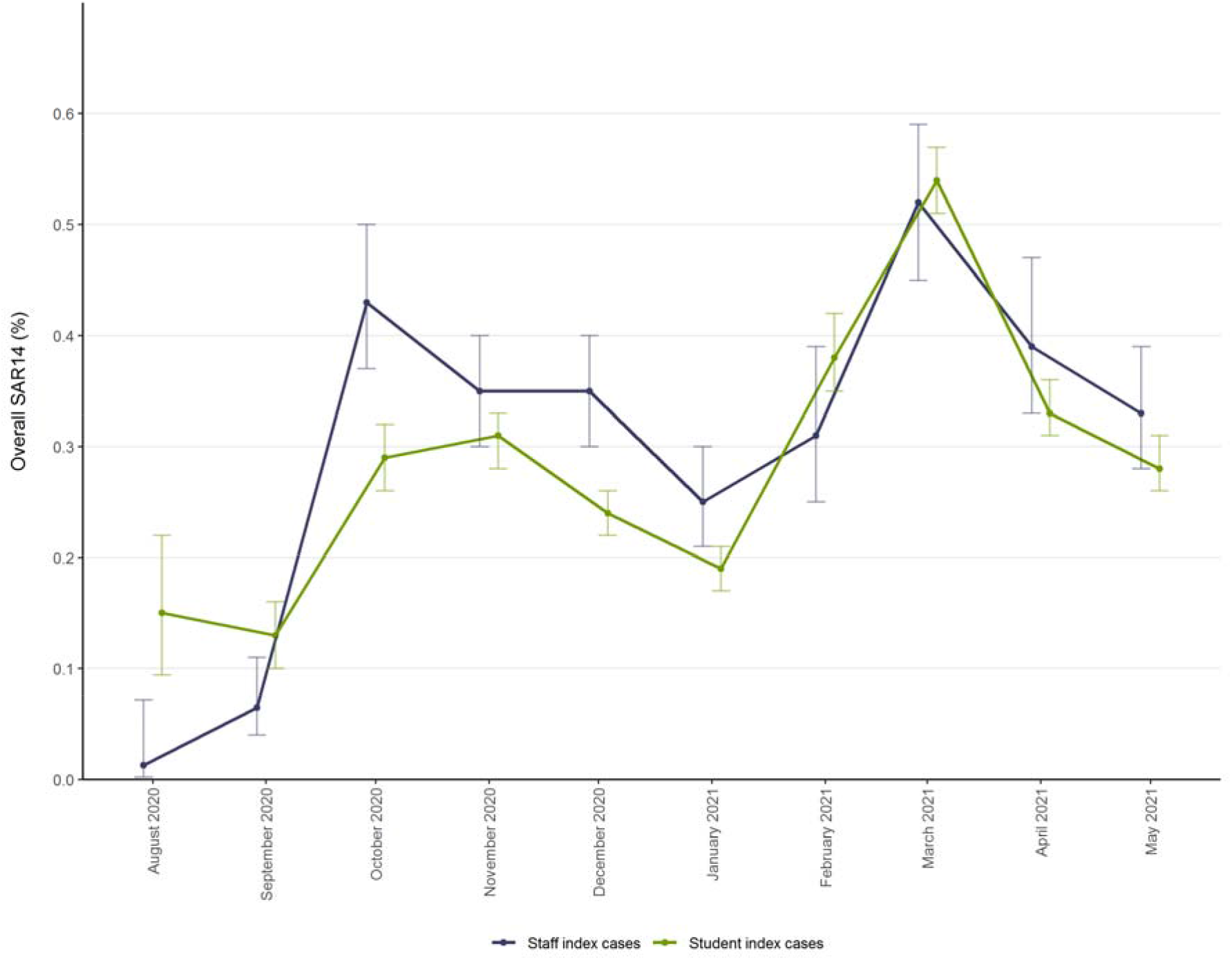
Secondary attack rates (95% CI), calculated by including both student and staff secondary cases infected within 14 days of index case, stratified by type of index case and month of positive SARS-CoV-2 test of index case.

The temporal trend in testing rates generally followed the trend in SAR (figure A-2), and approximately 10% of all students and 15% of all staff at a school were tested within 14 days after the index case tested positive (table A-4).

### Sensitivity analysis

The results from our sensitivity analysis, where we calculated SAR14 to students within each student birth cohort, did not notably differ from our main results on SAR14 to students (table A-5). The SAR14 ranged from 0.56% (95% CI 0.52-0.61) in students born in 2007 (grade 8) to 0.78% (95% CI 0.70-0.87) in students born in 2013 (grade 2).

## Discussion

Population-wide register data showed minimal transmission of SARS-CoV-2 between both students and staff in Norwegian primary and lower secondary schools during the academic year of 2020/2021. For most index cases, we observed no (55%) or only one (16%) secondary case.

Few students and educational staff at the school were infected in the 14 days following the positive test of the index case, measured by the secondary attack rate (SAR14). Less than 0.5 % of students and staff were infected within 14 days of the index case, regardless of characteristics of the index case. Throughout the academic year, transmission rates in schools largely followed the overall infection rates in the population, peaking in October/November 2020 and March 2021. However, the overall transmission in schools was low throughout the entire period, with SAR14 varying from just above 0% to 0.5%, even after the Alpha variant became dominant in Norway in February 2021 (17).

We observed a steady decline in SAR14 for both student and staff index cases following the first peak in October 2020. which could be explained by stricter IPC measures implemented as a response to high community infection rates. A similar decline in SAR14 was observed following the peak in March 2021, which was dominated by the Alpha variant. This further supports the effectiveness of implementing targeted IPC measures to reduce virus transmission in schools at times of high community incidence, also for more transmissible virus variants.

### Comparison with related studies

Previous studies have shown substantial variation in SARs of COVID-19 in different contact settings. Households have been the most important transmission venue, where meta-analyses have reported pooled SARs ranging from 16.4% to 20.0% (16, 18, 19). A Norwegian nation-wide study including all families with at least one parent and one child (comprising most students included in the current study) presented an overall SAR14 of 24 %, with even higher transmission rates when a parent rather than a child was the index case (1). In comparison, the school transmission rates presented in this study are exceptionally low, implying that transmission of COVID-19 is much more common within households and probably also in other social settings than within schools.

In contrast to our study, most existing studies on transmission of COVID-19 in schools have been conducted for selected schools or regions and within a shorter time frame, typically around the time of schools reopening for in-person learning after a period of school closure (3, 20-24). Our results support and advance those of existing studies, concluding that the importance of schools as an arena of transmission of COVID-19 is marginal when relevant IPC measures are in place (3). We observed similar findings in a setting where schools mainly were kept open during periods with rising community transmission.

The current study showed that transmission between students was slightly higher in lower secondary schools than in primary schools, supporting existing literature (25). Transmission from students to staff was higher in primary schools than in lower secondary schools. A possible explanation for this, is the closer contact between staff and younger children required for a secure and healthy psychosocial environment (26).

### Limitations of the study

To our knowledge, this is the first study of transmission rates of COVID-19 in schools using data representative for an entire nation, covering a full academic year. There are, however, certain limitations associated with our analysis. First, since transmission is far more likely to occur within the family than in school or other public domains (1, 15, 16), transmission between siblings attending the same school may have led to us overestimating SAR14. Lack of data on whether secondary cases were actually infected in school or elsewhere, may also have led to overestimation. However, if we are in fact overestimating SAR14, this would only *strengthen* our finding that Norwegian primary and lower secondary schools have not been important arenas for SARS-CoV-2 transmission.

Second, the registry data did not include information on which students and educational staff at the school were actual close contacts. Thus, we did not have the data necessary to estimate secondary attack rates among close contacts. Therefore, the aim of this study was not to estimate effects among close contacts, but to generate knowledge on general transmission in primary and lower secondary schools. The relatively high testing rates compared to transmission rates following the detection of an index case, indicate that most true secondary cases were captured in our data. Students in the same grade at the same school, are more likely to be close contacts than students at the same school in general, and all close contacts have routinely been tested as part of the standard IPC measures. Results from the sensitivity analysis on transmission between students in the same age cohort, were largely similar to the main results, which supports the robustness of the main results. Furthermore, all close contacts, usually involving the whole cohort or class, were urged to be tested if a positive case was identified at school (27). Modelling studies have shown that the Norwegian testing system has been well-functioning since the summer of 2020, with an estimated detection rate of more than 60% of all real cases (28).

Third, our data did not contain information on whether the index case had attended school or work during the infectious period. The policy in Norway of rapid testing, quarantining and isolation of infected cases and their close contacts, suggests that many index cases were in reality not present at school. Clearly, SAR14 is very likely to have been much higher if the IPC measures had not succeeded in quarantining suspected cases and keeping infected cases in isolation. However, this does not affect the main results of low risk of transmission in open schools with appropriate IPC policies in place. A smaller German study found an overall SAR in schools and day cares of 1.34% (24), which still supports our findings of low transmission rates in educational settings.

Lastly, the indirect identification of school affiliation based on small geographical neighbourhoods might results in some misclassifications of students to schools, though previous research suggests this is not a notable concern (29).

### Clinical implications and future directions

The findings of this study indicate that Norwegian primary and lower secondary schools have not been important arenas for COVID-19 transmission during the 2020/2021 academic year, despite being kept open. These findings add to the growing evidence that suggests it is safe to keep schools open during the pandemic, as long as appropriate IPC measures are implemented and adhered to (2, 3). In Norway, test-isolate-trace-and-quarantine has been the overarching strategy for limiting spread of SARS-CoV-2 (10). There have been societal IPC measures in place nationally throughout the study period, with further restrictions in areas with high community transmission. In addition, flexible IPC guidelines for schools were developed and implemented to reduce transmission (11, 12), as it has been a goal to avoid school closure due to its negative consequences for students’ learning and well-being (2, 6-9). The low transmission between students and staff observed in this study, indicates that these strategies have been appropriate to handle the situation in schools, also in areas with high infection rates after the Alpha variant became dominant. Of note, there was no masking policy in place for schools for most of the study period, questioning the need for extensive use of face masks in schools, given other IPC measures are implemented.

It might be that the implemented measures in open schools have been more effective in reducing overall virus transmission than school closure and digital education. Open schools allow students to meet and socialize in a controlled environment with IPC measures in place. Closed schools may, in addition to the mentioned negative consequences for learning and well-being, lead to less efficient contact tracing and delayed testing. Additionally, students who are not socializing in school, may over time find other arenas to socialize, possibly with higher transmission risk (1, 15, 16). Transmission in schools cannot be seen isolated from community transmission and combining school-targeted strategies with general IPC measures targeting the whole community, is crucial to limit transmission both into, within and outside schools.

Although our overall results showed very low transmission rates in schools, some variation by characteristics of the index case was observed. Transmission rates were found to be somewhat higher when a staff member, particularly teachers, was the index case. However, after the Alpha variant was introduced and eventually dominated, no significant difference could be observed between these groups. This may be explained by increased transmissibility of the Alpha variant across all age groups (30), including children (2). At the same time vaccination of adults in high-risk groups (but not students) was initiated, which could have had some significance for transmission rates involving staff members or for transmission rates from adult family members to students. The success of vaccination strategies will most likely be important for effective targeting of IPC measures in schools in the future.

Our data did only permit inclusion of primary and lower secondary schools. Higher infection rates of COVID-19 are generally observed among older, compared to younger, students (31). Whether this translates into higher transmission rates among students and educational staff in upper secondary schools remains unclear. Models made by the European Centre for Disease Prevention and Control (ECDC) show that closing secondary schools has larger effect on community transmission than closing primary schools or nurseries (2). However, as school closure should be avoided to the extent possible for all age groups, more knowledge on transmission in upper secondary schools is needed to help identify effective alternatives. Furthermore, future studies should address how vaccine coverage and more transmissible virus variants affects the transmission patterns in schools and households.

## Conclusion

SARS-CoV-2 did not spread to more than one additional individual in most cases where the virus was introduced in schools. Only a small proportion of all index cases led to clusters of three or more secondary infections, and transmission rates from students to school staff were generally very low. This suggests that primary and lower secondary schools, which were mainly kept open in Norway, were not important arenas for transmission of SARS-CoV-2, as long as appropriate IPC measures were in place.

## Data Availability

No additional data available. The datasets that support the findings of this study contain sensitive information and cannot be shared by the authors due to privacy laws. Individual-level data for research are generally available within Norway upon application conforming with strict regulations and procedures.

## Acknowledgements

We would like to thank our colleagues, especially Petter Elstrøm, at the Norwegian Institute of Public Health who have been part of the team handling the COVID-19 situation at educational institutions, for valuable discussion on the topic.

