## Supplemental files for "Transmission of SARS-CoV-2 in Norwegian schools: A population-wide register-based cohort study on characteristics of the index case and secondary attack rates"

### Appendix

#### Student school affiliation

The available data did not include individual information on student school affiliation. However, the organizational number of all primary and lower secondary schools were available for the vast majority of students who turned in the mandatory national tests in the academic year of 2019/2020. Also all student could be linked to their geographically defined neighborhood of residence (>12 000 in Norway), and students were then linked to schools based on their place of residence and birth year. Private schools are very uncommon in Norway, and students are attributed schools according to the school catchment areas, which are based on these geographically defined neighborhoods. It is also very uncommon in Norway to start school before or after the calendar year the child turns 6 years of age. Failing class is extremely rare and almost solely due to serious health conditions. Our approach is thus considered to yield reliable student-school matches (1).

#### Tables

**Table A-1. List of occupations included in the study population**

| Occupation | Code* |
| --- | --- |
| Teachers | 2341, 2342 |
| Child-care workers | 5311 |
| Teaching assistants | 5312 |

*Notes:*
Occupation was identified using the Employer- and Employee-register, as described in Magnusson et al. (2).
*According to the International Standards for Classification of Occupation (ISCO-08/STYRK-08)

**Table A-2. Distribution of index-cases across types of infection cluster (n (%)).**

|  | **Single case (n=2230)** | **Two-case clusters (n=631)** | **Multiple case clusters (n=1217)** | **Total (n=4078)** |
| --- | --- | --- | --- | --- |
| **School relation** |  |  |  |  |
| Students | 1708 (53.0) | 511 (15.9) | 1001 (31.1) | 3,220 (100.0) |
| Staff members | 522 (60.8) | 120 (14.0) | 216 (25.2) | 858 (100.0) |
| **School level** |  |  |  |  |
| Primary school | 1310 (56.0) | 367 (15.7) | 664 (28.4) | 2,341 (100.0) |
| Combined schools^a^ | 304 (54.7) | 78 (14.0) | 174 (31.3) | 556 (100.0) |
| Lower secondary schools | 616 (52.2) | 186 (15.7) | 379 (32.1) | 1,181 (100.0) |
| **Sex** |  |  |  |  |
| Female | 1208 (55.8) | 323 (14.9) | 635 (29.3) | 2,166 (100.0) |
| Male | 1022 (53.5) | 308 (16.1) | 582 (30.4) | 1,912 (100.0) |
| **School size** |  |  |  |  |
| < 100 | 152 (75.6) | 29 (14.4) | 20 (10.0) | 201 (100.0) |
| 100-299 | 715 (64.1) | 164 (14.7) | 236 (21.2) | 1,115 (100.0) |
| 300+ | 1363 (49.3) | 438 (15.9) | 961 (34.8) | 2,762 (100.0) |

*Notes:* ^a^Schools where primary and lower secondary school are combined

**Table A-3: Secondary attack rates (%) within 14 days after index test date (SAR14) stratified by demographic characteristics of index case.**

| **Characteristics of index case** | **Overall SAR14** | **SAR14 among students** | **SAR14 among school staff** |
| --- | --- | --- | --- |
| **All index cases** | **0.33 (5,816/1,784,982)** | **0.33 (5,089/1,555,139)** | **0.31 (727/230,843)** |
| Primary school | 0.32 (3,153/973,337) | 0.32 (2,710/847,431) | 0.35 (443/125,906) |
| Combined schools | 0.28 (840/300,475) | 0.28 (718/259,214) | 0.30 (122/41,261) |
| Lower secondary schools | 0.36 (1,823/511,711) | 0.37 (1,661/447,953) | 0.25 (162/63,758) |
| **County** |  |  |  |
| Oslo | 0.33 (1,340/408,765) | 0.31 (1,110/352,516) | 0.42 (230/55,249) |
| Rogaland | 0.21 (271/129,137) | 0.22 (245/113,692) | 0.17 (26/15,445) |
| Møre og Romsdal* | 0.11 (-) | 0.12 (-) | 0.038 (-) |
| Nordland | 0.25 (56/22,110) | 0.27 (51/18,757) | 0.15 (5/3,353) |
| Viken | 0.37 (2,396/647,188) | 0.38 (2,131/567,430) | 0.33 (265/79,758) |
| Innlandet | 0.36 (296/81,840) | 0.37 (260/69,963) | 0.30 (36/11,877) |
| Vestfold og Telemark | 0.35 (497/142,775) | 0.36 (447/124,501) | 0.27 (50/18,274) |
| Agder | 0.29 (250/87,805) | 0.29 (223/76,671) | 0.24 (27/11,134) |
| Vestland | 0.24 (438/172,783) | 0.25 (380/150,024) | 0.25 (58/22,759) |
| Trøndelag | 0.24 (174/77,255) | 0.23 (154/66,662) | 0.19 (20/10,593) |
| Troms og Finnmark | 0.23 (54/26,864) | 0.20 (46/23,242) | 0.22 (8/3,622) |
| **Sex** |  |  |  |
| Women | 0.32 (3,034/942,725) | 0.32 (2,640/820,244) | 0.32 (394/122,481) |
| Men | 0.33 (2,782/842,257) | 0.33 (2,449/733,895) | 0.31 (333/108,362) |
| **School size** |  |  |  |
| < 100 | 0.60 (92/15,256) | 0.69 (78/11,228) | 0.35 (14/4,028) |
| 100-299 | 0.36 (1,023/281,337) | 0.38 (899/239,469) | 0.30 (124/41,868) |
| 300+ | 0.32 (4,701/1,488,389) | 0.32 (4,112/1,303,442) | 0.32 (589/185,947) |
| **Students** | **0.32 (4,566/1,419,148)** | **0.33 (4,071/1,239,420)** | **0.28 (495/179,728)** |
| Primary school | 0.31 (2,665/859,332) | 0.31 (2,330/749,302) | 0.30 (335/110,030) |
| Lower secondary school | 0.34 (1,901/559,816) | 0.36 (1,741/490,118) | 0.23 (160/69,698) |
| **School staff** | **0.34 (1,250/365,834)** | **0.32 (1,018/314,719)** | **0.45 (232/51,115)** |
| Primary schools | 0.37 (813/219,525) | 0.35 (660/189,175) | 0.50 (153/30,350) |
| Combined schools | 0.26 (184/72,805) | 0.23 (145/62,039) | 0.36 (39/10,776) |
| Lower secondary schools | 0.34 (253/73,504) | 0.34 (213/63,505) | 0.40 (40/9,999) |
| Teacher | 0.37 (1,058/285,867) | 0.35 (864/246,442) | 0.49 (194/39,425) |
| Teaching assistants | 0.25 (124/50,329) | 0.24 (101/42,968) | 0.31 (23/7,361) |
| Child-care workers | 0.23 (68/29,638) | 0.21 (53/25,309) | 0.35 (15/4,329) |

*Notes:*
SAR14 was calculated using the number of infected non-index students and staff members in the numerator and the total number of non-index students and staff members in the denominator. This equation is given in parentheses (numerator/denominator) ^b^Schools where primary and lower secondary school are combined.
* Fewer than five secondary cases were censored due to privacy considerations.

**Table A-4: Percentage of students and staff tested within 14 days of index case.**

| **Characteristics of index case** | **% students and staff tested** | **% students tested** | **% staff tested** |
| --- | --- | --- | --- |
| **All index cases, all school levels** | **10.8 (10.8-10.9)** | **10.2 (10.2-10.3)** | **15.0 (14.9-15.2)** |
| **School size (all index cases)** |  |  |  |
| < 100 | 18.4 (17.8-19.0) | 18.4 (17.7-19.1) | 18.6 (17.4-19.8) |
| 100-299 | 13.1 (13.0-13.2) | 12.4 (12.2-12.5) | 17.3 (16.9-17.6) |
| 300+ | 10.3 (10.3-10.4) | 9.8 (9.7-9.8) | 14.4 (14.3-14.6) |
| **Students** | **11.0 (10.9-11.0)** | **10.5 (10.4-10.5)** | **14.7 (14.5-14.8)** |
| Primary school | 9.9 (9.8-9.9) | 9.3 (9.2-9.3) | 14.1 (13.9-14.3) |
| Lower secondary school | 12.7 (12.6-12.8) | 12.3 (12.2-12.4) | 5.6 (15.3-15.9) |
| **School staff** | **10.3 (10.2-10.4)** | **9.3 (9.2-9.4)** | **16.3 (15.9-16.6)** |
| Primary school | 9.9 (9.8-10.1) | 8.9 (8.8-9.0) | 16.5 (16.1-17.0) |
| Combined schools^b^ | 9.2 (8.9-9.4) | 8.2 (8.0-8.4) | 14.7 (14.0-15.4) |
| Lower secondary school | 12.3 (12.0-12.5) | 11.5 (11.3-11.8) | 17.1 (16.4-17.9) |
| Teacher | 10.7 (10.6-10.8) | 9.8 (9.6-9.9) | 16.8 (16.4-17.2) |
| Teaching assistants | 7.7 (10.6-10.8) | 6.7 (6.4-6.9) | 13.7 (12.9-14.5) |
| Child-care workers | 10.0 (9.7-10.4) | 9.0 (8.7-9.4) | 15.7 (14.7-16.8) |

*Notes:*
95 % confidence intervals in parentheses calculated using the Wilson method for binomial proportions. Test percentages were calculated as the number of non-index students and staff members who were tested for SARS-CoV-2 with a PCR test within 14 days of the confirmed COVID-19 index case, regardless of test result, divided by the total number of non-index students and staff members (analogously to the calculation of SAR14). Students or staff who had more than one test within 14 days are counted once.
 ^b^Schools where primary and lower secondary school are combined.

**Table A-5: Secondary attack rates (%) within 14 days after index test date (SAR14) by birth year.**

| **Characteristics of index case** | **SAR14 to students** | **Numerator/denominator** |
| --- | --- | --- |
| **School grade (birth year)** |  |  |
| 1 (2014) | 0.65 (0.62-0.72) | 283/43,351 |
| 2 (2013) | 0.78 (0.70-0.87) | 349/44,685 |
| 3 (2012) | 0.65 (0.59-0.73) | 313/47,827 |
| 4 (2011) | 0.69 (0.62-0.76) | 358/51,955 |
| 5 (2010) | 0.66 (0.59-0.73) | 337/51,435 |
| 6 (2009) | 0.68 (0.62-0.75) | 397/58,043 |
| 7 (2008) | 0.73 (0.66-0.80) | 413/56,964 |
| 8 (2007) | 0.56 (0.52-0.61) | 568/101,337 |
| 9 (2006) | 0.57 (0.53-0.62) | 615/107,805 |
| 10 (2005) | 0.67 (0.62-0.72) | 701/104,650 |
| *Notes:*  95 % confidence intervals in parentheses calculated using the Wilson method for binomial proportions. SAR14 was calculated using the number of infected non-index students in the numerator and the total number of non-index students in the denominator, stratified by birth year of the index case. ^b^Schools where primary and lower secondary school are combined. | | |

#### Figures

**Figure A-1. Percentage of total student (left) and school staff (right) population by county with confirmed COVID-19 throughout the academic year of 2020/2021.**

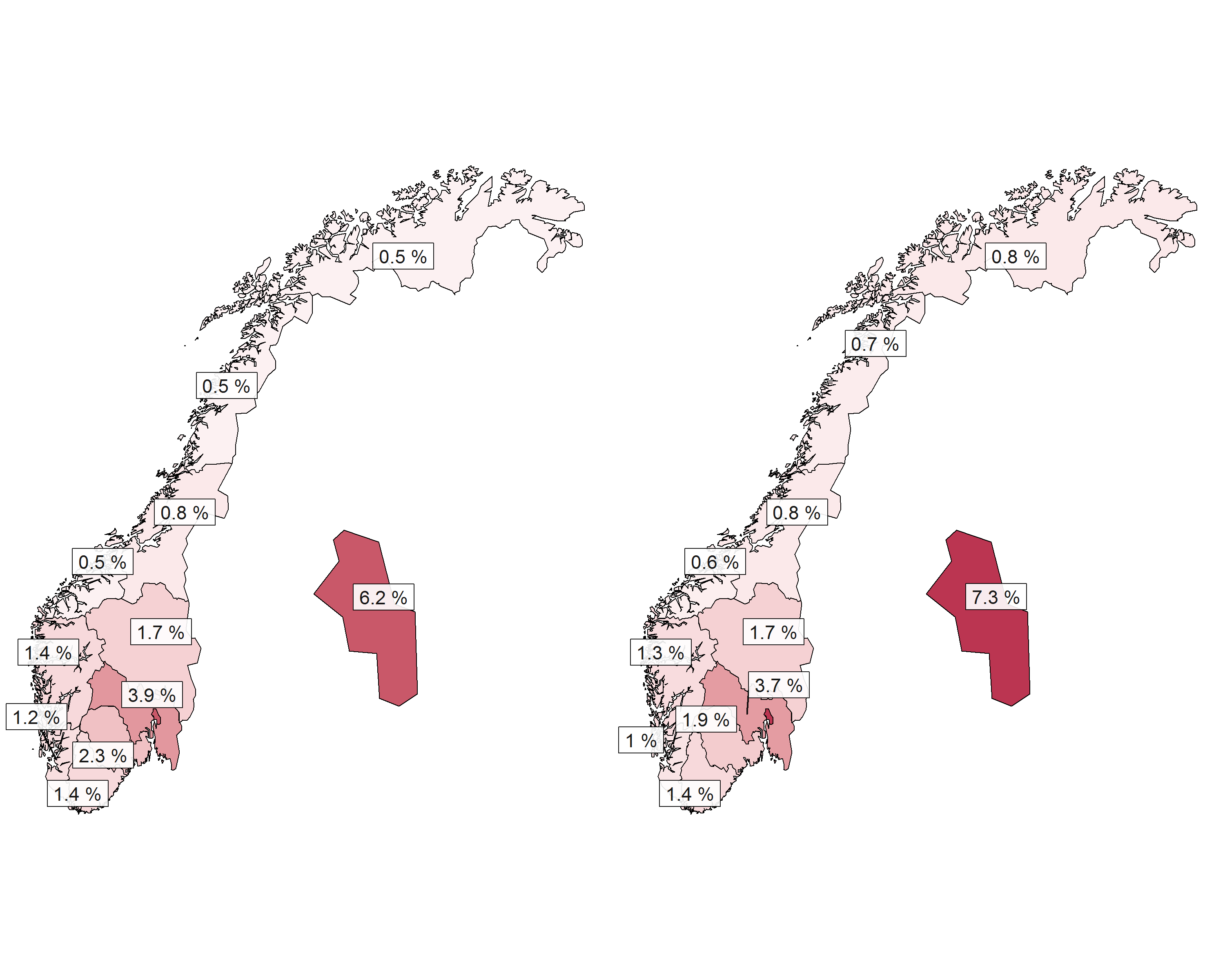

**Figure A-2. Secondary attack rates (SAR) and percentage of non-index students and staff tested for COVID-19 (secondary axis) within 14 days of index case positive.**

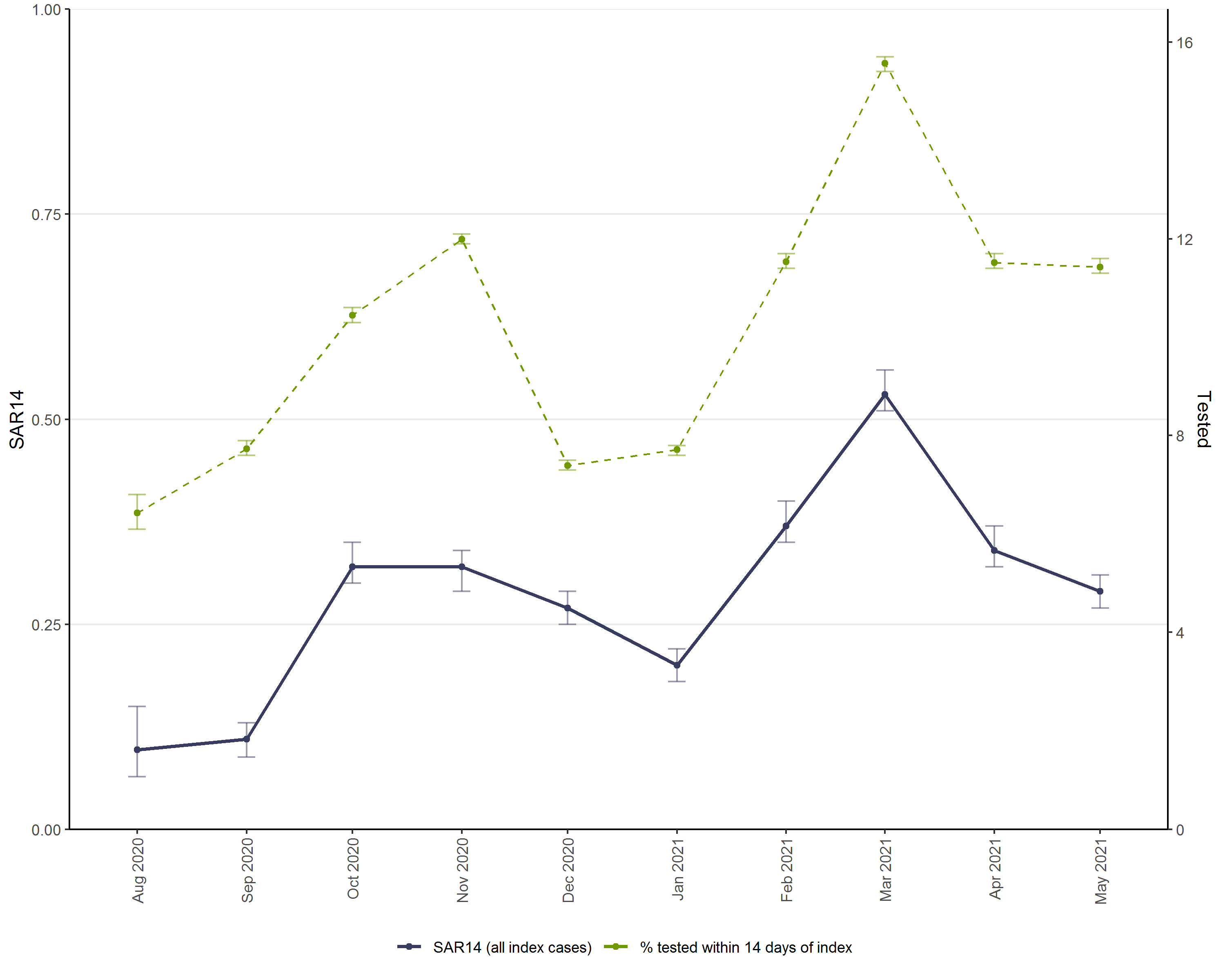

*Notes:*
SAR14 was calculated using the daily cumulative number of infected non-index students and staff members in the numerator and the total number of non-index students and staff members in the denominator. Test percentages were calculated by dividing the daily cumulative sum of tested students and staff by the total number of non-index students and staff. 95 % confidence intervals calculated using the Wilson method for binomial proportions.
